# Global shifts in thermal suitability and population at risk for dengue transmission by *Aedes spp.* mosquitoes under CMIP6 scenarios

**DOI:** 10.64898/2026.07.02.26357126

**Authors:** Sadie J. Ryan, Catherine A. Lippi, Leah R. Johnson, Jakob Meredith

## Abstract

Dengue fever risk and burden has increased globally in the past decade, with record-breaking outbreaks driving high case numbers, outbreaks increasing in existing transmission suitable regions, and occurring in new locations. A combination of global change processes, including climate change, have provided the environmental backdrop for introductions and resurgences of mosquito-transmitted dengue virus. Understanding shifts in exposure risk is integral to public health preparedness. This study provides global mapping of the thermal suitability of dengue transmission for CMIP6 climate scenarios, across a range of general circulation models (GCMs), and we created spatially explicit demographic projections of transmission risk using year-matched RCP-SSP frameworks for demographic and emissions scenarios. Globally, poleward shifts in projected distributions of suitability for transmission for both *Ae. aegypti* and *Ae. albopictus* suitability are shown in both the near term (2030s) and longer term (2050). Under a ‘middle of the road’ climate scenario (CMIP6 SSP2-4.5), regions in Africa and Asia are the major areas driving increases in year-round (12 months) population at risk (PAR) through 2050, with an anticipated net gain in 932 million people at risk for *Ae. aegypti* transmission and 24 million for *Ae. albopictus*, which includes multiple regions losing areas of year-round suitability as temperatures exceed the higher thermal boundary for transmission. In contrast, the estimated net increase in PAR for one or more months of transmission suitability at a global scale by 2050 is 3.29 billion people for *Ae. aegypti* transmission and 3.30 billion for *Ae. albopictus* transmission. This snapshot of a ‘middle-of-the-road’ combination of climate and demographic driven increases in potential dengue transmission exposure emphasizes the importance of both expanding suitability in new areas, and growing populations in areas approaching and becoming exposed year-round. Globalization, urbanization, and shipping will continue to provide the potential for introductions into newly suitable areas as season lengths increase, sparking outbreaks in unexposed populations. This is compounded and becomes ever more probable as the number of people and places at year-round risk also increases. This project provides all global gridded outputs for onward mapping and reuse, to add to the toolkit to anticipate and prepare for prevention and response to dengue in a changing world.

## Introduction

Dengue fever is caused by an orthoflaviviral infection by one of four serotypes of dengue virus (DENV 1-4), manifesting a range of symptoms and severity, from asymptomatic or mild flu-like symptoms to severe manifestations of dengue shock syndrome (DSS) and dengue hemorrhagic fever (DHF) [1]. Cases of dengue fever have been rising around the world, with outbreaks occurring with record case numbers (e.g. Nepal, Brazil), and occurring or recurring in new locations (e.g. France, California) [2–5]. In 2024, the World Health Organization’s global surveillance system reported over 14 million cases of dengue, which was double the number seen the year before and 12 times as many as in 2014 [2,6]. A combination of global change processes, including climate change [7], increased global travel and trade [8–10], and a relaxation of vector control activities in historical dengue hotspots due to a lack of economic or political investment [11] have provided the environmental backdrop for introductions and resurgences of *Aedes aegypti* and *Aedes albopictus* mosquito-transmitted dengue virus.

The mosquito vectors for dengue are now well adapted to human environments. *Ae. aegypti* is a competent vector for yellow fever, Zika virus, chikungunya, and many more human febrile diseases [12]. The spread of *Ae. aegypti* is facilitated by anthropophily - an evolved biting preference for humans and close association with human activity [13], and as a result has successfully exploited intercontinental invasion pathways driven by human movements [10,14,15]. It was likely introduced into the Americas from Africa multiple times, but significantly as a function of the slave trade [16,17], and has become well-established in much of North and South America. It is now reported to be present in 167 countries, and predicted to continue to spread into more tropical, subtropical, and temperate areas with little previous exposure [18]. *Ae. aegypti* is well adapted to the human built landscape and exploits urban and rapidly urbanizing environments around the world, taking advantage of water left in the environment, for larval habitat. It breeds in and around houses, planters, water storage containers, puddles, pockets in rubbish that can hold water, food containers, and non-residential areas such as construction sites, which can provide ample potential larval habitat - even independently of rainfall events [19]. *Ae. albopictus*, originally from Asia, has more recently also been introduced to new regions, and as a more temperate mosquito, has rapidly spread into regions not prepared for diseases previously assumed to be strictly ‘tropical’ [5,20]. In its native range, *Ae. albopictus* exploits a variety of natural and artificial habitats for oviposition, including bamboo stumps, tree holes, discarded tires, and catch basins [21]. In the US, *Ae. albopictus* is less suited to urban environments, and is instead associated with natural grassy or forested areas where there is ample larval habitat such as tree holes, making them a greater threat to communities at the edge of wooded areas or suburbs [22]. The role of *Ae. albopictus* spread in Europe in recent years is thought to be key to increasing reports of both dengue and chikungunya virus in the region [23,24].

In 2017, Mordecai et al. [25] developed and published a thermal suitability model for dengue transmission for both *Ae. aegypti* and *Ae. albopictus*, and Ryan et al. 2019 [26] used this model to project baseline and future predicted thermal suitability and population at risk (PAR) from future shifts in the thermal limits to suitability, using climate projections developed for the AR5 coupled model intercomparison project (CMIP5). The resultant output projections were subsequently also adapted and incorporated into the narrative for Chapter 12 on Central and South America of the IPCC6 report [27], among other re-uses of the publicly available gridded outputs, e.g. [28,29]. Since the development of these original projections in 2019, the CMIP6 climate scenarios have been developed and described in the 2023 IPCC report, and spatially explicit projections using the newer RCP-SSP (which we will refer to SSP in this paper) framework have been developed and made publicly accessible (e.g. WorldClim.org [30]).

This study provides updated global mapping of the thermal suitability of dengue transmission by *Ae. aegypti* and *Ae. albopictus* using CMIP6 products. While there are many modeling approaches to climate-driven dengue prediction and transmission [31–35], the goal here is only to provide an update on gridded output, and a summary of CMIP6 driven outcomes, through the lens of populations at risk (PAR). An important aspect of our use of updated climate models is to frame them in context of a concern raised in response to the AR6 ensemble used in the IPCC6 scenario projections, which is that some of the models were thought to predict higher rates and levels of global warming than expected, perhaps leading to overly pessimistic projections in the ensemble model used [36]. We thus take an approach of using not an ensemble but a range of outcomes from 5 general circulation models (GCMs), which balance two key sensitivity metrics within the range of ‘likely’ global warming outcomes; this is detailed in the methods section.

In this study, we present updated gridded climate-driven dengue transmission suitability outcomes using a different but overlapping suite of GCMs than in prior work (2019, 2023). We generated outcomes for 2021-2040, referred to as 2030, and 2041-2060, referred to as 2050, across five GCMs (see methods), under two scenarios of greenhouse gas emissions, SSP 2-4.5 and SSP 5-8.5, but we focus on describing RCP2-4.5, a “middle of the road” scenario, in this paper. These are projected from a baseline period of 1970-2000 [30], which we refer to as baseline. The projections are mapped to explore the shifting geography of the number of months suitable for transmission into 2030 and 2050, and we explore the impact of these shifts through the lens of time-matched projected populations at risk (PAR) across global regions.

## Methods and Materials

### Thermal Transmission Suitability Models

Thermal boundaries for dengue transmission by the two mosquito vectors, *Aedes aegypti* and *Aedes albopictus* are described in [26]. The thermal boundaries for dengue transmission by *Ae. aegypti* and *Ae. albopictus* used here were originally calculated using a mechanistic modelling approach, fitting thermal performance curves for each component in the transmission life cycle to derive an overall *R_0_* curve as a function of temperature [25]. Subsequently, Ryan et al. [26] evaluated the probability that *R_0_* > 0 at each 0.1℃ temperature, for a conservative posterior probability Prob(*R_0_* > 0) = 0.975 to derive dengue transmission suitability (S(T)) as a function of temperature for each species [26]. The thermal suitability bounds for dengue transmission by *Ae. aegypti* were 21.3°C and 34°C, and 19.9°C and 29.4°C for *Ae. albopictus* [26].

### Climate Data

Baseline (1970-2000) monthly mean temperature data were obtained from WorldClim.org [30]. For downscaled future projected climate we selected five global circulation models (GCMs) for the period we refer to as 2030 (2021-2040), and 2050 (2041-2060), for CMIP6 SSP 2-4.5 and SSP 5-8.5. All climate data were downloaded at a 30s resolution.

To account for an issue in CMIP6 models used in the AR6 IPCC reporting, known colloquially as “the hot model issue”, wherein several GCMs included in the AR6 ensemble predicted larger temperature increases, and may have introduced a hotter bias into interpreted description of impact [36], we chose to assess and select GCMs for our projected climate scenarios as follows. We examined two key sensitivity metrics for the individual GCMs, transient climate response (TCR), and equilibrium climate sensitivity (ECS), and chose a set of GCMs spanning a range of sensitivities. Specifically, following the logic presented in [36], wherein TCR =1.4-2.2 falls in the ‘likely’ range, and ECS=2.5-4 falls in the ‘likely’ range, we assigned GCMs with metrics within the range as medium (M), and thus values of TCR and ECS outside these ranges are high (H) or low (L). We chose five GCMs to demonstrate the range, as shown in Table 1, consistent with methodology in [37].

**Table 1.** The five general circulation models (GCMs) selected. (abbreviations (ABB) and full model names (GCM)) to illustrate the range of model outcomes within the CMIP6 suite, using measures of model sensitivity (transient climate response, TCR, and equilibrium climate sensitivity, ECS), designated Low (L), Medium (M) and High (H) to rank ‘hottest’ to ‘coldest’ projections (see methods). (adapted from [37])

| ABB | GCM | ECS (°C) | TCR (°C) | Ranking |
| --- | --- | --- | --- | --- |
| HAD | HadGEM3-GC31-LL | 5.55 | 2.55 | HH (hottest) |
| ACC | ACCESS-CM2 | 4.72 | 2.10 | HM |
| CMC | CMCC-ESM2 | 3.55 | 1.92 | MM |
| MRI | MRI-ESM2-0 | 3.15 | 1.64 | ML |
| MIR | MIROC6 | 2.61 | 1.55 | LL (coldest) |

### Population Data

For population data and demographic projections, we used the approach described in projecting Zika transmission exposures in Ryan et al 2021 [38], which deviates from the Ryan et al. 2019 [26] methodology, in that future population at risk (PAR) estimates are based on matched Shared Socioeconomic Pathway (SSP) projections [39,40], instead of an assumption of constant population size. This responds to the 2019 paper, which pointed to the unavailability of the projected population grids at that time. Additionally, this paper updates the SSP projections to the 2020 release of the ‘revision 01 dataset’ [39], which enables regional population calculations on grids with higher resolution pixels. Importantly, the terminology for CMIP6 models shifted from CMIP5 to include SSPs explicitly, so the correct matching underlying population projections are relevant here, that is the SSP2 and SSP5 projections. For our baseline population, we used the baseline year 2000, based on the Gridded Population of the World, v4 [41], and the baseline used for the projected SSP population products here [39]. Note that this sets our baseline year back an additional 15 years in contrast to the 2019 paper. These gridded population data were downloaded from NASA Earth Data (https://search.earthdata.nasa.gov/) at their 0.125 degree resolution. Lastly, we subdivided the global populations and corresponding suitability into 21 geographic and socioeconomic regions as defined by the Global Burden of Disease (GBD) studies [42]. We shortened these region names for visualisation purposes, and full and shortened names are given in Table S1. Regionalized PAR was calculated using a modified version of the R package ‘fasterize’ [43], R package ‘terra’ [44], and a study specific version of our generalized script pipeline for mapping thermal suitability as a function of climate data layers found at: https://github.com/RyanLab/stephensimaps [45].

## Results

The baseline pattern of predicted thermal suitability for dengue transmission aligns with the expected pattern for viruses transmitted by *Ae. aegypti* and *Ae. albopictus*, where much of the tropics are suitable for year-round transmission (Figs. 1 and 2, panels A and B). Dengue transmission suitability is expected to shift geographically in the future for both mosquito species under all projected combinations of climate models and development scenarios. There is a marked poleward expansion of transmission suitability seen for both mosquito species, where previously unsuitable locations on the periphery of baseline ranges become marginally suitable for dengue transmission into the future (Figs. 1 and 2). This includes marginal northward expansion of suitability into Canada, Central Europe, and Russia, which is more pronounced for *Ae. albopictus* (Figs. 1 and 2 panel D). Although poleward gains in suitability are less pronounced for *Ae. aegypti*, future extension of the transmission season is seen across ‘cooler’ (Fig. 1C) and ‘hotter’ (Fig. 2C) climate models, particularly along mid-latitude regions that border the subtropics at baseline. In contrast, some reduction in the length of dengue transmission seasons is expected to occur with *Ae. albopictus* throughout the tropics, and these reductions are more extensive under the ‘hotter’ climate model, as temperatures exceed the maximum suitability (Fig. 2D).

**Fig. 1.**
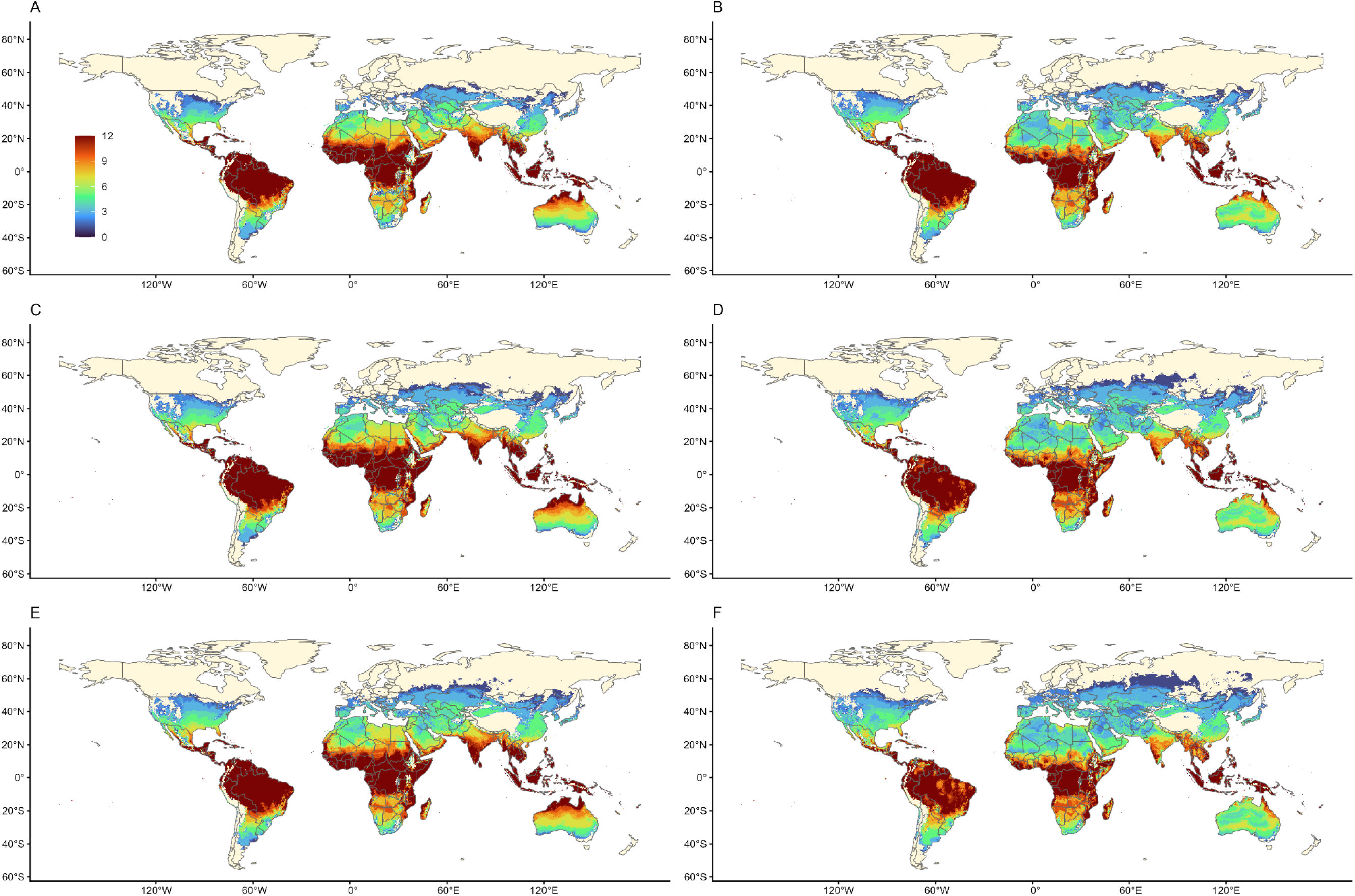
Number of months (0-12) of suitability for dengue transmission (S(T)>0, with 97.5% probability), for *Aedes aegypti* (A, C, E) and *Aedes albopictus* (B, D, F) at baseline (A, B), 2030 (C,D) and 2050 (E,F) for the MIR GCM (‘coolest’ See Table 1), for CMIP6 SSP 2-4.5. Maps made using Natural Earth Data in R [46]

**Fig. 2.**
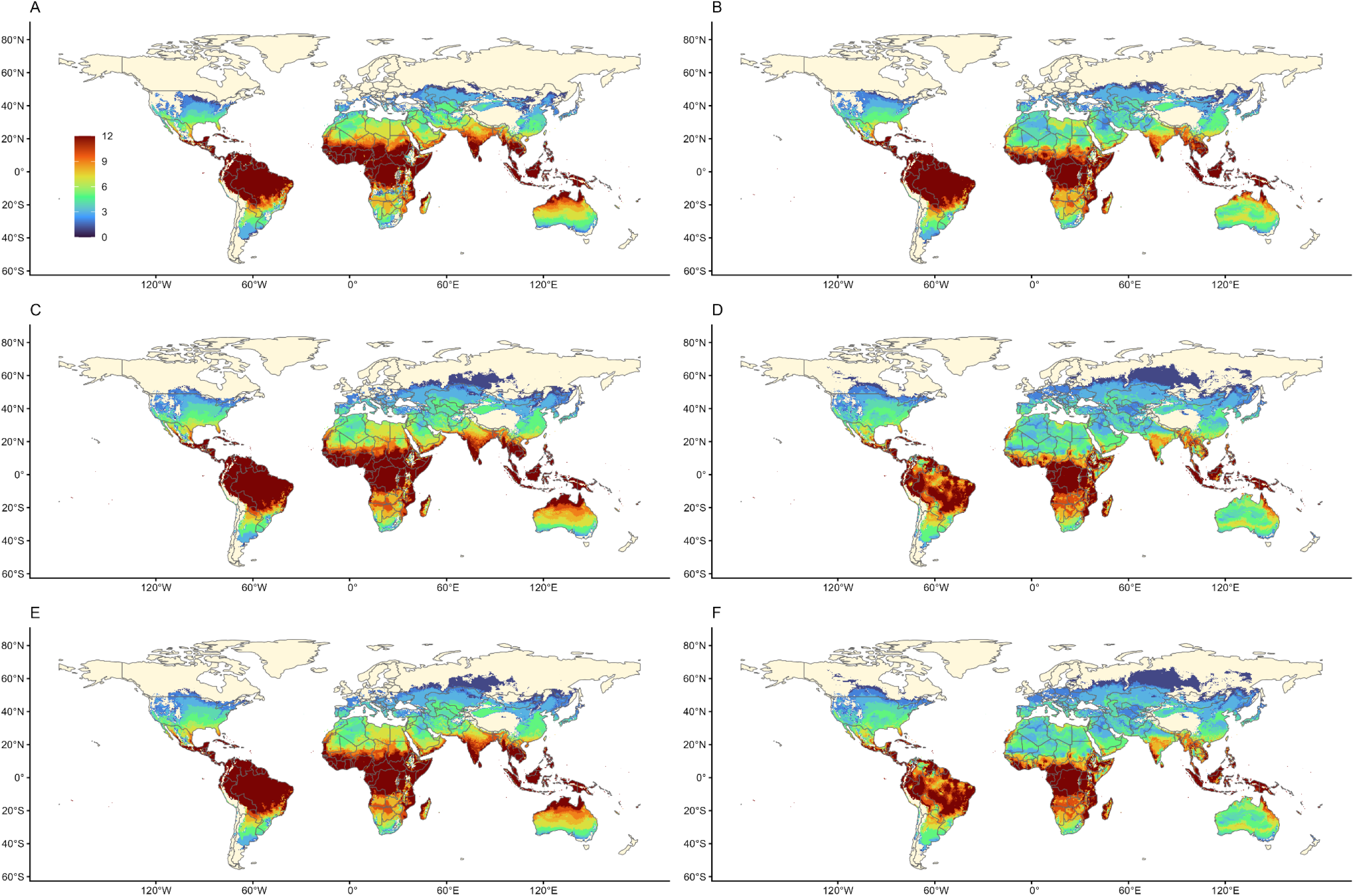
Number of months of suitability for dengue transmission (S(T)>0, with 97.5% probability), for *Aedes aegypti* (A, C, E) and *Aedes albopictus* (B, D, F) at baseline (A, B), 2030 (C,D) and 2050 (E,F) for the HAD GCM (‘hottest’ See Table 1), for CMIP6 SSP 2-4.5. Maps made using Natural Earth Data in R [46]

Geographic shifts in transmission suitability are accompanied by changes in the projected population at risk of exposure to dengue (PAR). We illustrate these changes for the middle-road scenario SSP2-4.5, and in terms of one or more months (Figure 3) or year-round (12 months) exposure (Figure 4). Figure S1 depicts maps of the most extreme scenario combination generated (HAD, SSP5-8.5, 2050) in contrast to baseline, for both *Ae. aegypti* and *Ae. albopictus* transmission, as an illustration of the unlikely nature of SSP5-8.5 and the ‘hottest’ GCM.

**Fig 3.**
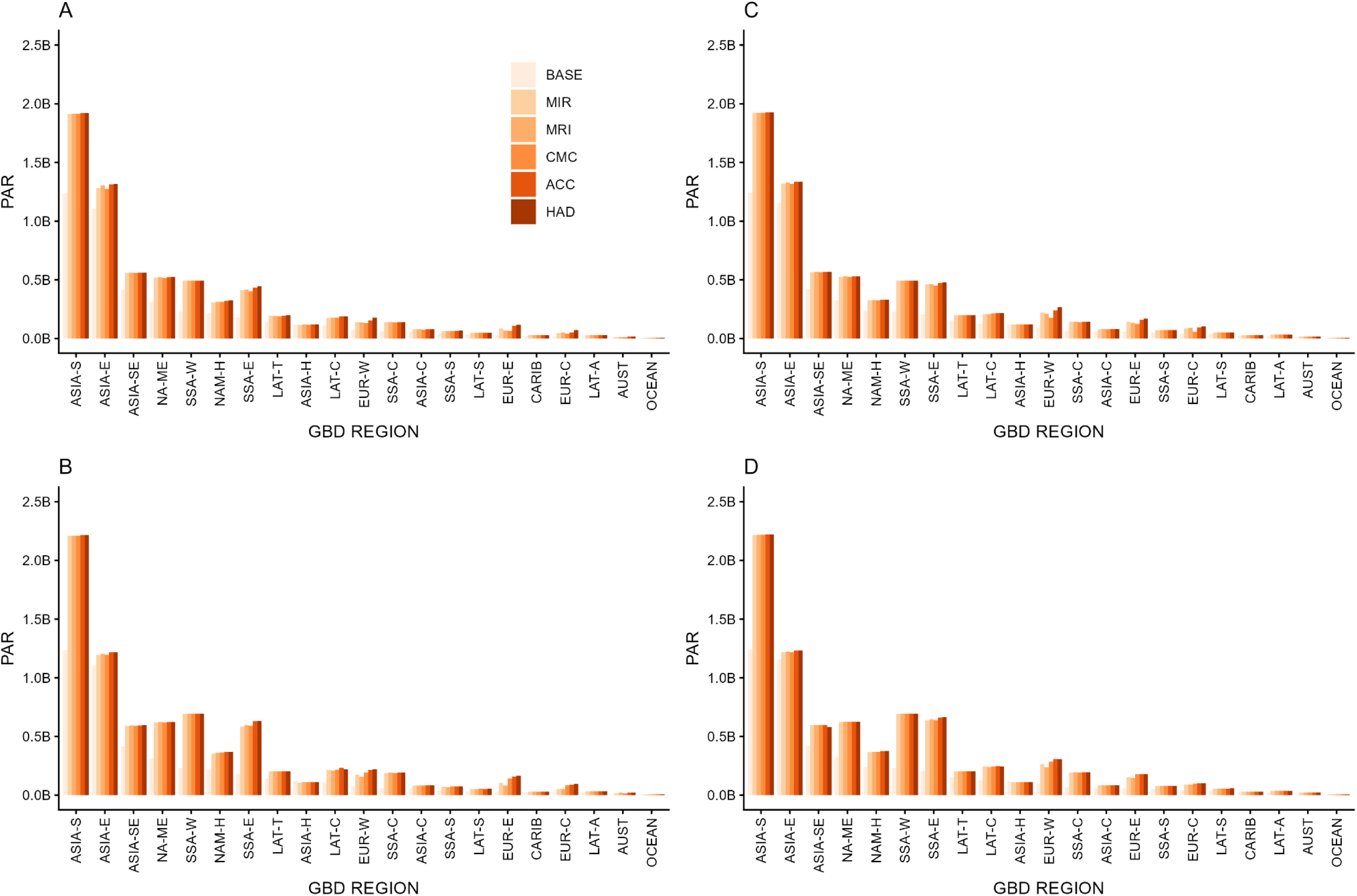
population at risk (PAR) for one or more months of dengue transmission suitability for *Aedes aegypti* (A, B) and *Aedes albopictus* (C, D) in 2030 (A, C) and 2050 (B, D) by GCM for SSP2-4.5, including baseline (BASE) PAR (for full tabulated results and GBD region names, See Tables 1–4). Note that regions are ordered by BASE PAR, which differs by *Aedes* species.

**Fig 4.**
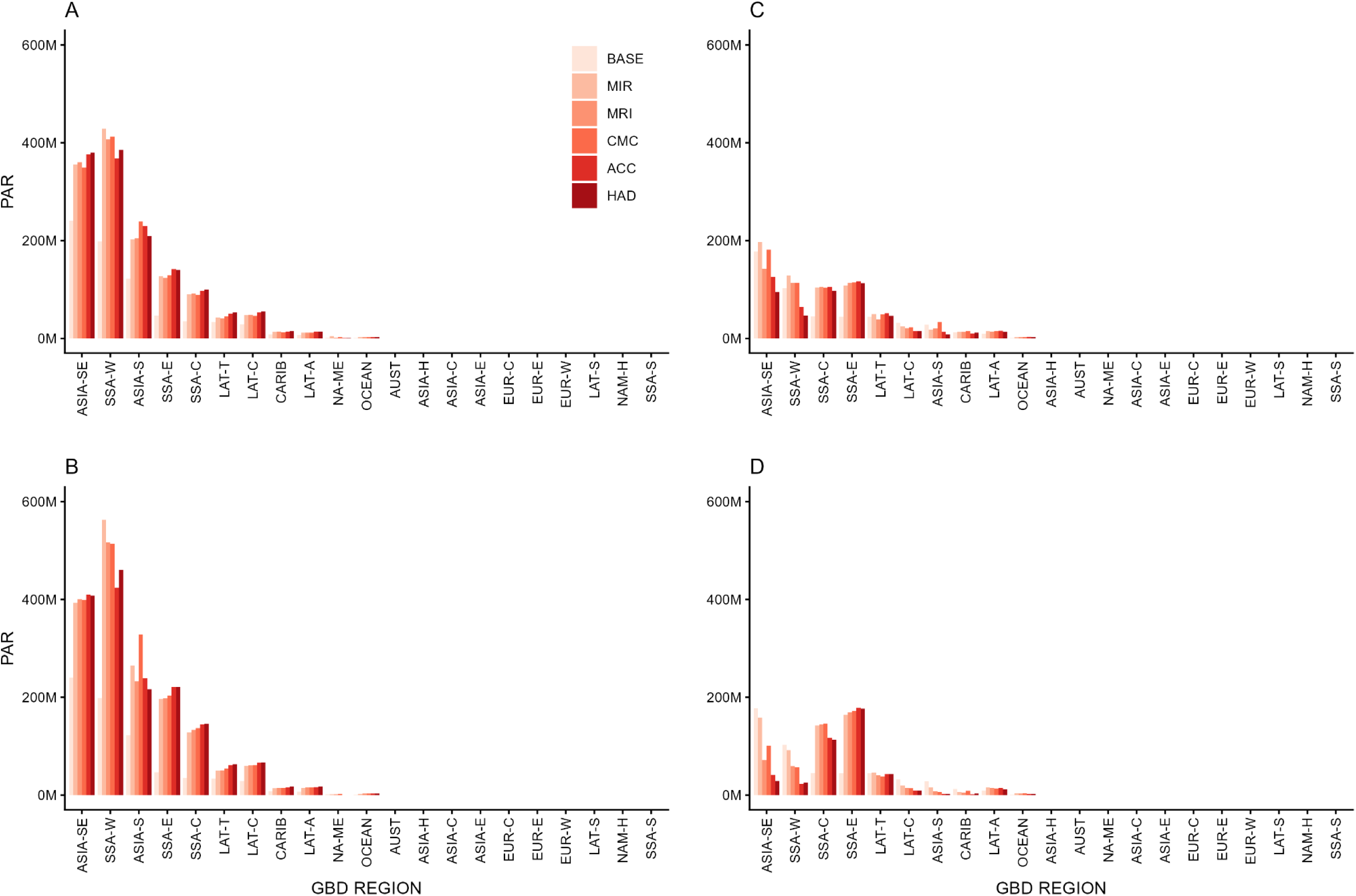
population at risk (PAR) year-round for dengue transmission suitability for *Aedes aegypti* (A, B) and *Aedes albopictus* (C, D) in 2030 (A, C) and 2050 (B, D) by GCM for SSP2-4.5, including baseline (BASE) PAR (for full tabulated results and GBD region names, See Tables 1–4). Note that regions are ordered by BASE PAR, which differs by *Aedes* species.

The highest projected change in numbers for PAR is for one or more months of dengue transmission suitability, as expected. The South and East Asia regions are expected to have the most PAR, for both mosquito species, across all climate models through 2050 (Fig. 3). With the exception of the High-Income Pacific Asia region, every region is projected to experience a net gain in PAR for at least one month of dengue transmission suitability (Table 2). This reflects the increase in geographic area for even low numbers of months of suitability (Figs. 1 and 2), overlaid on the projected growth of the population itself across different regions (Table S2). Note that this is a middle-road (SSP2-4.5) scenario, and using extreme scenarios for geographic shifts (Figure S1) would likely generate far larger estimates of PAR. The estimated net increase in PAR for one or more months of transmission suitability at a global scale by 2050 is 3.29 billion people for *Ae. aegypti* transmission and 3.30 billion for *Ae. albopictus* transmission.

**Table 2:** Increase in estimated population at risk (PAR) from 2000 to 2050 under SSP 2-45 for *Aedes aegypti* (A) and *Aedes albopictus* (B) for one or more months of dengue transmission suitability, taking the mean across our five GCMs. Global Burden of Disease (GBD) Regions ordered from highest to lowest PAR; decreased PAR indicated by (parentheses).

| Regions (ordered) | New PAR (*Aedes aegypti*) | Regions (ordered) | New PAR (*Ae albopictus*) |
| --- | --- | --- | --- |
| Asia (South) | 976,670,479 | Asia (South) | 976,848,349 |
| Sub-Saharan Africa (West) | 465,714,236 | Sub-Saharan Africa (West) | 465,579,724 |
| Sub-Saharan Africa (East) | 429,028,178 | Sub-Saharan Africa (East) | 443,804,802 |
| North Africa & Middle East | 311,098,239 | North Africa & Middle East | 302,766,342 |
| Asia (Southeast) | 176,363,230 | Europe (Western) | 186,115,088 |
| North America (High Income) | 147,105,058 | Asia (Southeast) | 171,178,329 |
| Sub-Saharan Africa (Central) | 129,901,432 | North America (High Income) | 131,707,319 |
| Europe (Western) | 115,780,104 | Sub-Saharan Africa (Central) | 128,985,290 |
| Latin America (Central) | 105,293,470 | Latin America (Central) | 114,861,420 |
| Europe (Eastern) | 101,132,581 | Europe (Eastern) | 109,266,336 |
| Asia (East) | 100,254,652 | Asia (East) | 66,829,252 |
| Latin America (Tropical) | 64,321,504 | Europe (Central) | 55,830,375 |
| Europe (Central) | 54,743,260 | Latin America (Tropical) | 53,682,613 |
| Sub-Saharan Africa (Southern) | 34,624,808 | Sub-Saharan Africa (Southern) | 25,651,186 |
| Asia (Central) | 28,777,004 | Asia (Central) | 25,295,806 |
| Latin America (Southern) | 19,581,714 | Latin America (Southern) | 17,910,976 |
| Latin America (Andean) | 14,891,173 | Latin America (Andean) | 14,534,544 |
| Australasia | 12,047,150 | Australasia | 11,965,237 |
| Caribbean | 5,753,198 | Caribbean | 5,322,376 |
| Oceania | 4,678,744 | Oceania | 5,102,459 |
| Asia (High Income Pacific) | (6,462,319) | Asia (High Income Pacific) | (9,783,270) |
| **Net Gains** | **3,291,297,896** |  | **3,303,454,555** |

By the year 2030, we expect to see increases in the year-round PAR of dengue exposure by *Ae. aegypti* across 9 GBD regions under all climate models, which will continue to increase by 2050. The largest increases in PAR of year-round dengue exposure from *Ae. aegypti* will be in Southeast Asia, western sub-Saharan Africa, southern Asia, and eastern sub-Saharan Africa (Fig. 4A, 4B). In contrast, many regions will see smaller increases in PAR of year round dengue exposure from *Ae. albopictus* (Fig. 4C, 4D). By the year 2050, some regions including southeast Asia, western sub-Saharan Africa, and central Latin America are expected to have decreases in PAR as increasing temperatures in these regions become too hot to support year-round transmission (Fig. 4D).

In total, the estimated net increase in year-round PAR through 2050 is 932.4 million for *Ae. aegypti*, and 24.1 million for *Ae. albopictus* transmission suitability (Table 3). *Ae. aegypti* is only expected to have future reductions in year-round PAR in North Africa and the Middle East, and the large net gains in PAR are driven by increases in both areas suitable for transmission and the length of transmission seasons. Despite considerable reductions in transmission suitability and PAR throughout many regions (Table 3), overall increases in transmission suitability for *Ae. albopictus* are expected to drive net increases in PAR that will outpace any localized decreases.

**Table 3:** Increase in estimated population at risk (PAR) from 2000 to 2050 under SSP 2-45 for *Aedes aegypti* (A) and *Aedes albopictus* (B) for year-round dengue transmission suitability, taking the mean across our five GCMs. Global Burden of Disease (GBD) Regions ordered from highest to lowest PAR; decreased PAR indicated by (parentheses).

| Regions (ordered) | New PAR (*Aedes aegypti*) | Regions (ordered) | New PAR (*Ae albopictus*) |
| --- | --- | --- | --- |
| Sub-Saharan Africa (West) | 296,989,752 | Sub-Saharan Africa (East) | 127,366,948 |
| Asia (Southeast) | 161,780,283 | Sub-Saharan Africa (Central) | 87,272,648 |
| Sub-Saharan Africa (East) | 161,309,102 | Latin America (Andean) | 4,556,807 |
| Asia (South) | 134,353,198 | Oceania | 2,152,825 |
| Sub-Saharan Africa (Central) | 102,849,317 | Sub-Saharan Africa (Southern) | 40,260 |
| Latin America (Central) | 34,216,431 | North Africa & Middle East | 1,201 |
| Latin America (Tropical) | 22,556,203 | Asia (Central) | - |
| Latin America (Andean) | 8,590,646 | Asia (East) | - |
| Caribbean | 7,714,013 | Europe (Central) | - |
| Oceania | 2,205,369 | Europe (Eastern) | - |
| North America (High Income) | 45,291 | Europe (Western) | - |
| Asia (High Income Pacific) | 35,150 | Latin America (Southern) | - |
| Australasia | 34,689 | North America (High Income) | - |
| Sub-Saharan Africa (Southern) | 10,879 | Australasia | (8,779) |
| Asia (East) | 5,114 | Asia (High Income Pacific) | (13,077) |
| Asia (Central) | - | Latin America (Tropical) | (1,815,519) |
| Europe (Central) | - | Caribbean | (7,118,898) |
| Europe (Eastern) | - | Latin America (Central) | (18,399,156) |
| Europe (Western) | - | Asia (South) | (21,221,942) |
| Latin America (Southern) | - | Sub-Saharan Africa (West) | (51,181,699) |
| North Africa & Middle East | (281,508) | Asia (Southeast) | (97,502,819) |
| **Net Gain** | **932,413,928** |  | **24,128,799** |

In this paper we tabulated summaries and charted information for the SSP 2-4.5 scenario, to highlight a middle-road future with changing climate and population growth. We additionally modeled SSP 5-8.5 scenarios, and all global gridded model outputs (n=42) are available through Harvard Dataverse (https://dataverse.harvard.edu/dataverse/DEN_CMIP6).

## Discussion

In this study, we provide an overview of projected geographic and demographic changes in dengue thermal suitability of transmission from 2000 to 2030 and 2050 using the CMIP6 scenarios framework. Echoing findings for CMIP5 scenarios, we see poleward shifts in suitability for both *Ae. aegypti* and *Ae. albopictus* in the near (2030s) and longer term (2050) projections (Figures 1,2). This general pattern holds true across the ‘coolest’ and ‘hottest’ GCMs (Table 1) in this study. However, differences in the impact of the choice of GCM can be seen in the degree of poleward shifts (for example, along the Canada border and into Northern Europe), and the degree to which suitability decreases, in terms of the number of months suitable, in Brazil, for example, for *Ae. albopictus* transmission, as predicted temperatures exceed suitability maximum in some months in 2030 and 2050 (Figures 1 and 2). This provides an update to previous work [26], which presented a striking view of potential geographic shifts and impacts under the most optimistic (RCP 2.6) and most pessimistic (RCP 8.5) projections of climate. In this study, we describe a ‘middle of the road’ SSP2-4.5 view instead, but we also provide gridded output estimates for both CMIP6 SSP2-4.5 and SSP5-8.5 across a suite of GCMs for 2030 and 2050 for onward reuse (https://dataverse.harvard.edu/dataverse/DEN_CMIP6).

Recent work emphasizes a necessary shift away from RCP8.5/SSP 5-8.5 framings as we move into the new cycle of CMIP7 [47], as they represent an overly extreme and unlikely future scenario (see Fig S1). Our ‘middle of the road’ SSP2-4.5 approach nonetheless found that an estimated 3.29 billion additional people are projected to be newly at risk for dengue transmission suitability by *Ae. aegypti* 2050, and 3.30 billion by *Ae. albopictus*.

The SSP framework provides a means to incorporate differential projected demographic responses at a global scale, under the different socioeconomic pathways prescribed. This is reflected in both regional underlying demographic trajectories in this study, and the global scale differences in projected population growth. In our example scenario, summing across our GBD regions for this gridded SSP population product [39], the overall global population changes from 5.46 billion in 2000 to 8.33 billion under SSP2-4.5 in 2050. In an apparent paradox, this is higher than the summed population under SSP5-8.5 for 2050, of 7.74 billion, as the population in more extreme SSPs is anticipated to peak and decline earlier in the century. In order to describe the potential impact of shifting geography of dengue transmission suitability under future climate scenarios, we calculated the population at risk (PAR) of both one or more months of suitability, and year-round suitability (12 months). To remain concise, we show PAR in raw numbers, as projected, in Figures 3 and 4, under the SSP 2-4.5 scenario in 2050, across 21 regions [42]. Figure 3 shows largest net gains in PAR in Asian and SSA regions, with some re-ordering of regional impact by mosquito, also shown in Table 2, wherein *Ae. albopictus* transmission shows more gains in the Latin American Andean region and Oceania than other regions. Figure 4, whose population axis is roughly one third that of Figure 3, highlights the impact of increased endemic (12 month) risk anticipated in Asian and sub-Saharan African (SSA) regions, and to a smaller but important degree, tropical Latin America. This depiction of impact incorporating the SSP projected demographies creates a striking regional distribution of PAR, and these changes are distributed differently across the regions (Table S2), leading to large regional increases in risk amplified by demographic shifts in regions of geographic shifts. In the previous study [26], regions in Europe contributed greatly to projected PAR totals, topping the list of individual regions contributing to overall PAR increases in 2080. The previous study described outcomes under the CMIP5 RCP8.5 scenario which predicted dramatic poleward shifts in risk (see 2019 Figure 2B) and a much larger geographic distribution of exposure to that risk, comparable to our Figure S1C and D, posing a striking contrast to our current use of SSP 2-4.5. While this study generates less extreme estimates of population at risk (PAR) than the previous study, we nonetheless find that 3.29 billion additional people are estimated to be at risk from dengue transmission suitability (one or more months) from *Ae. aegypti*, and 3.30 billion from *Ae. albopictus* by 2050 compared to baseline. Our results also place regions in Africa and Asia as the major areas driving increases in year-round (12 months) PAR through 2050, with regions in Europe accounting for none of the net changes (Table 2), indicating that under this middle-road climate scenario, endemic dengue risk concentrates in regions of high population growth. However, if we look at areas experiencing one or more months of risk, we see gains in every region, except high income Asia (Table 3). This suggests that we will see increased outbreaks in new regions, as new areas are exposed to suitable temperatures for more of the year, and season length will increase in non-endemic areas.

Incorporating the thermal suitability limits of each vector species into models underlies the development of biologically meaningful projections and risk assessments. Explicitly accounting for the role that temperature plays in determining broad environmental suitability is important, given that nearly every aspect of mosquito life history and pathogen transmission is heavily dependent on temperature. That said, in the environment the thermal limits of mosquito populations may not be quite as rigid as implemented in this work. *Ae. aegypti* has some capacity for local physiological thermal adaptation [48], as well as behavioral shifts, such as microhabitat selection of cooler areas [49], that may determine habitat and thus transmission suitability on localized scales. Further, temperature is not the only factor that determines habitat or transmission suitability in mosquitoes. Although our global projections are limited to temperature suitability, other factors such as humidity also contribute to whether mosquitoes can successfully occupy an environment [50]. However, the empirical evidence necessary to begin to fully incorporate these factors into models for these species of mosquitoes are not currently available. There is also evidence to suggest that thermal adaptation of the vector may not alter viral and bacterial temperature response [51]. Thus, despite these limitations, these thermal boundaries reflect biological constraints on mosquitoes and can describe general patterns of thermal transmission suitability.

## Conclusion

Globalization, urbanization, and international trade will continue to provide potential introduction pathways for dengue vectors into newly suitable areas, sparking outbreaks in unexposed human populations as transmission seasons increase in length. This study provides an updated snapshot of a ‘middle-of-the-road’ scenario, combining climate and demographic driven increases in potential dengue transmission exposure. This work emphasizes the importance of both expanding suitability in new locations and growing populations, as the number of people and places at year-round risk of dengue exposure also increases. Providing updated estimates of potential shifts in dengue exposure is important to ensure that public health planning and policy decisions are made with the best information possible. By producing outputs that leverage best available climate products and newer development scenarios, we are providing a resource to support the uptake of newer climate information into modeling and policy. To that end, this project provides all global gridded outputs (https://dataverse.harvard.edu/dataverse/DEN_CMIP6) for onward mapping and reuse, to add to the toolkit to anticipate and prepare for prevention and response to dengue in a changing world.

## Data Availability

All global gridded model outputs (n=42) are available through Harvard Dataverse (https://dataverse.harvard.edu/dataverse/DEN_CMIP6)

## Supporting Information

S1 Table: Global burden of disease (GBD) regions (n=21) full and shortened names

S2 Table: Population by GBD region (n=21) for 2000 and projected SSPs in 2030 and 2050 for SSP2-4.5 and SSP5-8.5

S1 Figure: Number of months of suitability for dengue transmission (S(T)>0, with 97.5% probability), for *Aedes aegypti* (A, C) and *Aedes albopictus* (B, D) at baseline (A, B), and 2050 (C, D) for the HAD GCM (hottest’ See Table 1), for CMIP6 RCP 5-8.5. Maps made using Natural Earth Data in R [46]

## Acknowledgments

This work was supported by the Centers for Disease Control (CDC) Cooperative Agreement Numbers 1U01CK000510 and 1U01CK000662: Southeastern Regional Center of Excellence in Vector-Borne Diseases: The Gateway Program. Its contents are solely the responsibility of the authors and do not necessarily represent the official views of the Centers for Disease Control and Prevention or the Department of Health and Human Services. SJR, CAL and LRJ were supported by NSF DBI 2016265 and NSF DBI 2016264 as part of CIBR: VectorByte: A Global Informatics Platform for studying the Ecology of Vector-Borne Diseases. SJR would like to thank Jaye Madden for all the work and admin support at UF throughout this project.

---

**Table S1:** Global burden of disease (GBD) regions (n=21) full and shortened names.

| REGION | ABB |
| --- | --- |
| Asia (Central) | ASIA-C |
| Asia (East) | ASIA-E |
| Asia (High Income Pacific) | ASIA-H |
| Asia (South) | ASIA-S |
| Asia (Southeast) | ASIA-SE |
| Australasia | AUST |
| Caribbean | CARIB |
| Europe (Central) | EUR-C |
| Europe (Eastern) | EUR-E |
| Europe (Western) | EUR-W |
| Latin America (Andean) | LAT-A |
| Latin America (Central) | LAT-C |
| Latin America (Southern) | LAT-S |
| Latin America (Tropical) | LAT-T |
| North Africa & Middle East | NA-ME |
| North America (High Income) | NAM-H |
| Oceania | OCEAN |
| Sub-Saharan Africa (Central) | SSA-C |
| Sub-Saharan Africa (East) | SSA-E |
| Sub-Saharan Africa (Southern) | SSA-S |
| Sub-Saharan Africa (West) | SSA-W |

**Table S2:** Population by GBD region (n=21) for 2000 and projected SSPs in 2030 and 2050 for SSP2-4.5 and SSP5-8.5.

| REGION | 2000 | 2030 SSP2-4.5 | 2050 SSP2-4.5 | 2030 SSP5-8.5 | 2050 SSP5-8.5 |
| --- | --- | --- | --- | --- | --- |
| Asia (Central) | 72,278,661 | 87,376,568 | 90,698,317 | 82,062,488 | 77,900,754 |
| Asia (East) | 1,239,761,220 | 1,364,337,772 | 1,247,631,614 | 1,343,431,347 | 1,210,407,001 |
| Asia (High Income Pacific) | 122,673,029 | 121,335,551 | 110,087,552 | 126,037,835 | 123,583,592 |
| Asia (South) | 1,264,048,437 | 1,944,285,238 | 2,236,849,417 | 1,840,739,180 | 1,961,003,345 |
| Asia (Southeast) | 429,755,285 | 570,924,658 | 600,689,883 | 551,165,861 | 552,119,019 |
| Australasia | 13,300,609 | 19,857,458 | 24,056,267 | 21,744,439 | 29,666,136 |
| Caribbean | 25,352,929 | 30,138,974 | 29,950,176 | 28,038,230 | 25,180,777 |
| Europe (Central) | 115,469,941 | 111,707,994 | 105,817,528 | 114,214,980 | 113,522,038 |
| Europe (Eastern) | 205,550,209 | 189,440,086 | 182,730,751 | 190,218,365 | 183,795,623 |
| Europe (Western) | 325,906,922 | 367,260,210 | 383,539,100 | 388,605,157 | 447,908,799 |
| Latin America (Andean) | 39,917,646 | 52,496,572 | 56,293,939 | 48,192,274 | 46,291,762 |
| Latin America (Central) | 185,948,077 | 262,641,159 | 291,211,371 | 246,153,396 | 248,548,307 |
| Latin America (Southern) | 49,339,806 | 60,094,392 | 63,531,795 | 57,893,991 | 57,841,587 |
| Latin America (Tropical) | 153,863,409 | 197,875,008 | 206,550,337 | 190,611,811 | 189,002,446 |
| North Africa & Middle East | 342,360,298 | 534,464,726 | 629,418,961 | 510,822,043 | 564,379,095 |
| North America (High Income) | 264,803,696 | 338,853,580 | 377,733,695 | 362,529,415 | 450,282,158 |
| Oceania | 4,633,447 | 8,762,036 | 10,430,161 | 8,258,712 | 9,047,985 |
| Sub-Saharan Africa (Central) | 69,113,124 | 147,301,036 | 199,268,935 | 139,470,086 | 177,249,957 |
| Sub-Saharan Africa (East) | 253,700,240 | 525,521,854 | 709,145,069 | 489,068,603 | 598,750,565 |
| Sub-Saharan Africa (Southern) | 57,206,083 | 74,786,721 | 79,409,726 | 74,074,631 | 77,328,850 |
| Sub-Saharan Africa (West) | 229,679,226 | 493,025,828 | 695,214,239 | 460,479,954 | 591,297,197 |
| **GLOBAL** | **5,464,662,294** | **7,502,487,421** | **8,330,258,833** | **7,273,812,798** | **7,735,106,993** |

**Supplemental Figure 1:**
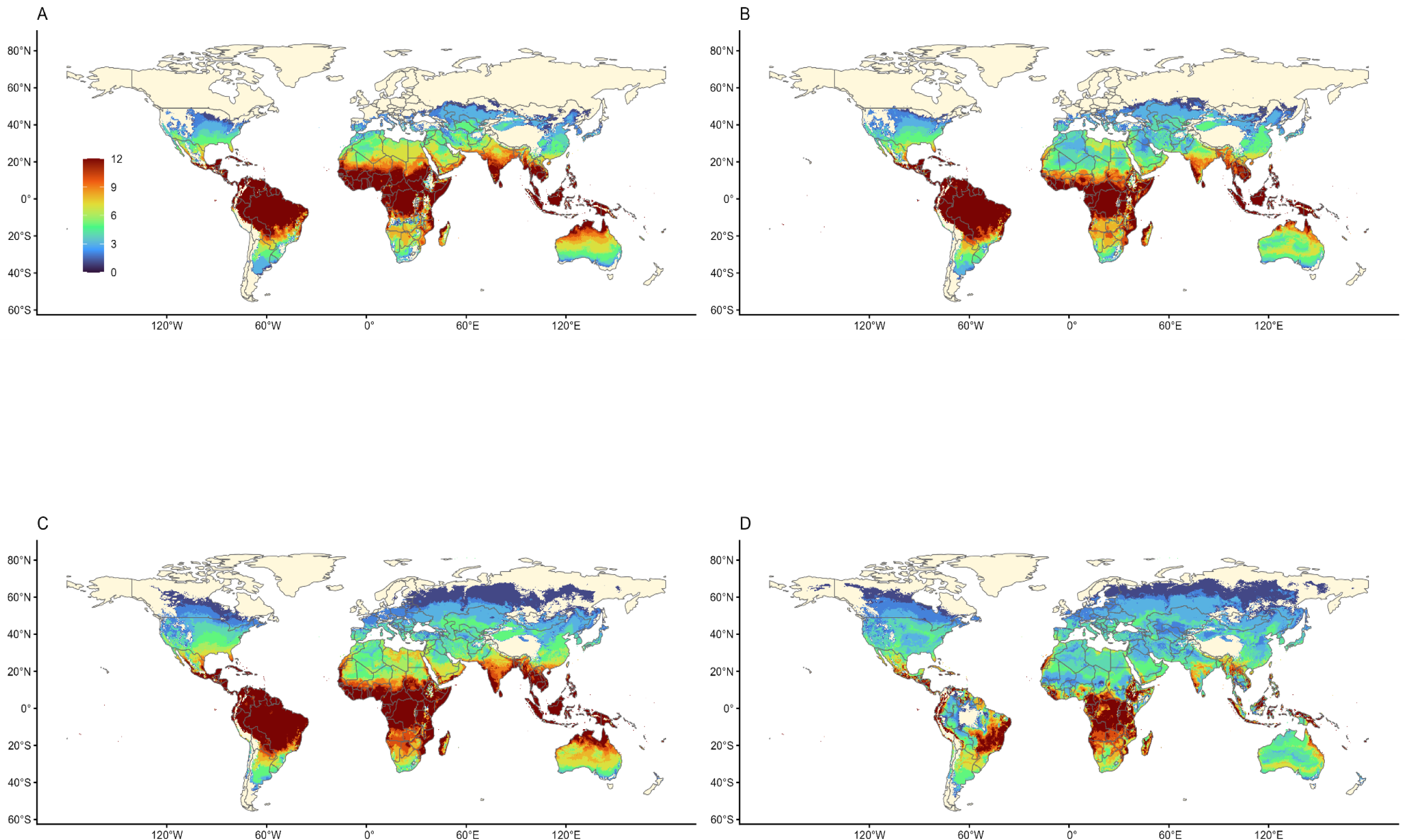
Number of months of suitability for dengue transmission (S(T)>0, with 97.5% probability), for *Aedes aegypti* (A, C) and *Aedes albopictus* (B, D) at baseline (A, B), and 2050 (C,D) for the HAD GCM (‘hottest’ See Table 1), for CMIP6 RCP 5-8.5. Maps made using Natural Earth Data in R [46]

